# Optimisation of whole cell human depletion provides increased sensitivity and microbial genome coverage from respiratory metagenomic assays

**DOI:** 10.1101/2025.10.10.25337716

**Authors:** Ronan M Doyle, Vishwa Patel, Gul Humayun, Renee Chang, Maria Gumbanmarasigan, Viktorija Vorslava, Benenteta Leka, Sophia Girgis, Dan Ward, Laura Truesdale, Julianne R Brown, Robbie Hammond, Angelika Kopec, Simon A Rock, Manoj Valappil, Kerry Laws, Surendra Parmar, Sophie May, Poorvi Patel, Muktar Ibrahim, Steve Green, Kordo Saeed, Nicholas Norton, Paul Grant, Gaia Nebbia, Rahul Batra, Jonathan Edgeworth, Luke B Snell, Adela Alcolea-Medina

## Abstract

Metagenomics is being adopted worldwide as a laboratory developed test. We have previously reported a unified metagenomic method for the direct detection of microorganisms from respiratory samples, delivering preliminary results within six hours with >90% sensitivity for both viral and bacterial pathogens. This has now been deployed across the United Kingdom (UK) National Health Service (NHS) through the NHS Genomic Networks of Excellence programme. Since this deployment we have identified a number of improvements to the method to further increase sensitivity. In this manuscript we report these optimisations, including the addition of a bead-beating cycle, substitution of the endonuclease enzyme, and controlled termination of the endonuclease reaction. This updated protocol (v1.2) resulted in substantially improved assay performance. Validation in six NHS hospitals using both protocols in parallel (v1 and v1.2) with 107 clinical respiratory samples demonstrated increased bacterial and viral read recovery in v1.2 translating to higher pathogen genome coverage compared to v1. Overall bacterial sensitivity for v1.2 was 98% compared to 93% for v1, and viral sensitivity was 90% for v1.2 compared to 80% for v1. This multicentre validation has confirmed reproducibility and scalability of the new protocol, establishing a robust framework for clinical implementation of metagenomics.

## Introduction

Metagenomics is a powerful method that enables genetic analysis of microbial communities in their natural environment. Due to the methods’ potential for complete pathogen characterisation in a single test, the adoption of metagenomics has gained traction as a clinical diagnostic method worldwide^1–4^. It has been applied to a range of sample types, including cerebrospinal fluid, tissues, and respiratory samples. However, existing methods often have prolonged turnaround times due to the labour-intensive nature of library preparation and extended sequencing duration. A major contributing factor to these delays is the high proportion of human DNA in clinical samples, which, if not depleted before processing, significantly impacts efficiency of pathogen genome recovery^5^.

We recently developed and implemented a unified metagenomic approach capable of detecting and sequencing a broad spectrum of pathogens in a single assay^6^. This includes bacteria, DNA and RNA viruses, fungi, and fastidious organisms such as *Mycoplasma, Treponema*, and *Chlamydia*, as well as parasites such as *Toxoplasma*. This method integrates quality control throughout the workflow, making it suitable for clinical laboratory implementation. Notably, 94% of results were reported within six and seven hours from sample to production of first actionable sequence report. It achieved a, achieving a sensitivity >90% and specificity of 99% after 24 hours of sequencing, as validated against standard clinical laboratory methods. The method was applied in the intensive care unit (ICU) at St Thomas’ Hospital (UK National Health Service [NHS]) during the winter of 2023–24 in a cohort of 74 patients. The results directly influenced clinical decision-making by intensive care physicians, leading to antimicrobial escalation or de-escalation and, with negative findings, the initiation of immunosuppressive therapy when a non-infectious inflammatory pathology was clinically suspected^7^.

To deplete human nucleic acids in clinical samples, the original protocol employed Heat-Labile Salt-Active Nucleases (HL-SAN, ArcticZymes Technologies). However, HL-SAN requires high concentrations of NaCl for optimal activity, which can compromise the integrity of microbial nucleic acids. To address this limitation, we optimised the method by replacing HL-SAN with Medium-Salt Active Nucleases (M-SAN, ArcticZymes Technologies), which operates efficiently at lower salt concentrations. We hypothesized that this modification would enhance pathogen detection sensitivity and genome recovery by improving human DNA/RNA depletion while preserving microbial nucleic acids.

Following the manufacturer’s recommendations, in the updated protocol magnesium ions (Mg^2^□) were also added to enhance M-SAN activity. In the updated protocol the reaction is now subsequently halted by the addition of EDTA, chelating the divalent cations and preventing unwanted degradation of microbial nucleic acids during downstream DNA/RNA extraction steps.

In addition to the enzymatic changes, we modified the mechanical lysis step. Samples were subjected to two rounds of bead-beating, rather than one, with the aim of increasing disruption of human cells and thereby improving depletion efficiency. Furthermore, for bronchoalveolar lavage (BAL) samples, the initial centrifugation step was omitted with the hypothesis of allowing better access of the enzyme to host cells.

This updated protocol was validated across six NHS hospitals and is now implemented nationally under the NHS Genomic Networks of Excellence programme. Sensitivity, specificity, and limit of detection were systematically assessed. The results of this multicentre validation study of the updated protocol are presented here.

## Results

### Intact microorganisms are preserved during the human depletion step

We assessed the impact of the human DNA depletion step on intact micro-organisms using control material containing intact viral and atypical bacterial targets [Respiratory Panel 2.1 (RP2.1) Controls (Zeptometrix™)]. Controls were processed using the previously validated human depletion step (protocol v1), the updated human depletion step (protocol v1.2), and a third condition in which extraction was performed without depletion. All three extracts were subjected to PCR targeting a panel of respiratory pathogens to assess the effects of depletion on intact pathogens. The cycle threshold (Ct) values across the three conditions showed minimal variation, indicating that the depletion steps in protocols v1 and v1.2 did not deplete intact viral or atypical bacterial targets (Table 1).

**Table 1.** Quantitative PCR Ct values of organisms in RP2.1 control sample after no human depletion, protocol v1 and protocol v1.2.

| Organism | Without human depletion | Protocol v1 | Protocol v1.2 |
| --- | --- | --- | --- |
| Influenza A | 29.4 | 29.3 | 28.8 |
| Influenza B | 26.6 | 27.0 | 27.1 |
| Respiratory Syncytial Virus | 31.5 | 31.1 | 31.7 |
| Parainfluenza 1 | 37.5 | 37.2 | 37.6 |
| Parainfluenza 3 | 39.1 | 38.7 | 37.9 |
| Parainfluenza 2 | 33.3 | 32.7 | 32.9 |
| Parainfluenza 4 | 35.3 | 35.3 | 35.7 |
| Human adenovirus | 28.9 | 27.2 | 29.7 |
| Metapneumovirus | 32.6 | 32.5 | 32.5 |
| Rhinovirus | 34.3 | 34.4 | 34.5 |
| Human coronavirus 229E | 31.0 | 30.3 | 30.9 |
| Human coronavirus OC43 | 35.0 | 34.0 | 35.0 |
| Human coronavirus HKU1 | 32.7 | 32.2 | 32.8 |
| *Chlamydia pneumoniae* | 33.3 | 31.4 | 33.6 |
| *Mycoplasma pneumoniae* | 34.4 | 32.5 | 34.2 |

### Increased human depletion in protocol v1.2

We compared human read depletion in 107 pairs of respiratory clinical samples in total, using the v1 protocol or v1.2 protocol in parallel across six separate sites (Site 1 n = 11, Site 2 n = 27, Site 3 n = 34, Site 4 n = 12, Site 5 n = 11 and Site 6 n = 12). Within the 107 sample pairs, six separate respiratory sample types were tested (BAL n = 40, ETT n = 37, NPA n = 6, NTS n = 7, PFL n = 2 and SPT n = 15). The total number of reads recovered after sequencing for both the v1 and v1.2 samples were comparable, with no statistically significant differences between either group (v1 median total reads = 133,000, v1.2 median total reads = 166,000, *p =* 0.511), nor for any of the different sites or sample types (Supplementary figures 1-3). However, there was both a statistically significant decrease in both the raw number and percentage of reads classified as human (Fig. 1, Supplementary figures 6 and 7) in v1.2 samples compared to their v1 pair. This translated to a median reduction in human reads of −28,800 (v1 median human reads = 74,300, v1.2 median human reads = 45,500, *p* = 0.002) or a decrease in relative abundance of human reads of –38% (v1 human relative abundance = 84%, v1.2 median human relative abundance = 46%, *p* = 1.42 × 10^−13^). A comparison of the same results split by sample type showed the greatest reduction in percentage of human reads in bronchoalveolar lavage (BAL) and nose and throat swabs (NTS) samples (Δ = −41%, v1 median relative abundance = 84%, v2 median relative abundance = 43%, *p* = 8.4 × 10^−7^ and Δ = −59%, v1 median relative abundance = 67%, v2 median relative abundance = 8%, *p* = 0.022, respectively) (Supplementary Figure 7). Endotracheal tube (ETT) and nasopharyngeal aspirate (NPA) samples also had a statistically significant decrease in the relative abundance of human reads prepared with v1 compared to v1.2 (Δ = −10%, p = 1.2 ×10^−5^ and Δ = −25%, *p* = 0.036, respectively) (Supplementary figure 7).

**Fig. 1.**
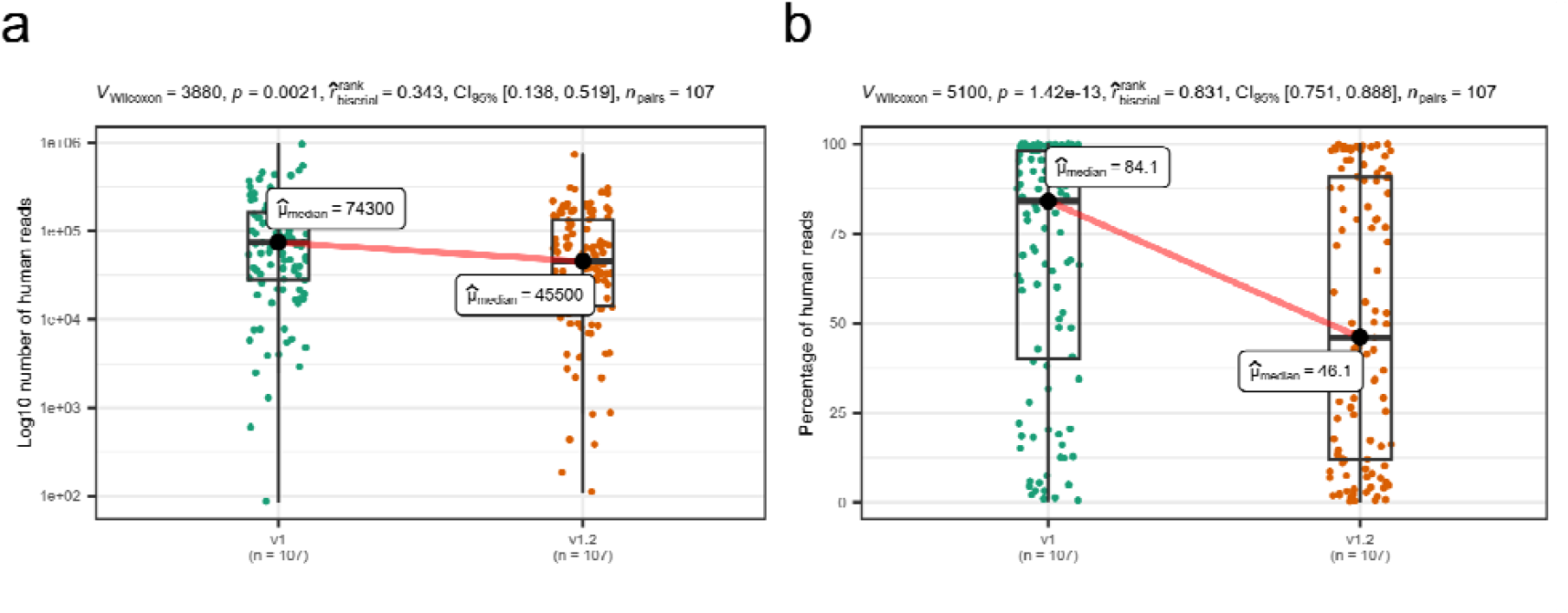
Boxplots displaying the difference between 107 samples prepared by either protocol v1 or protocol v1.2 for a) the Log10 total number of human reads classified in each sample. b) The percentage of reads per sample classified as human. Results from a Wilcoxon signed-rank test comparing the two groups is displayed at the top of the plot.

To confirm human depletion, we also tested a subset of 12 pairs of samples included in the above analysis by quantitative PCR targeting human DNA. There was a statistically significant reduction in amplified human DNA from v1.2 compared to v1, represented by a median increase in Ct value of +13.4 (v1 median Ct value = 26.6, v1.2 median Ct value = 34.0, *p* = 0.002) (Fig. 2).

**Fig. 2.**
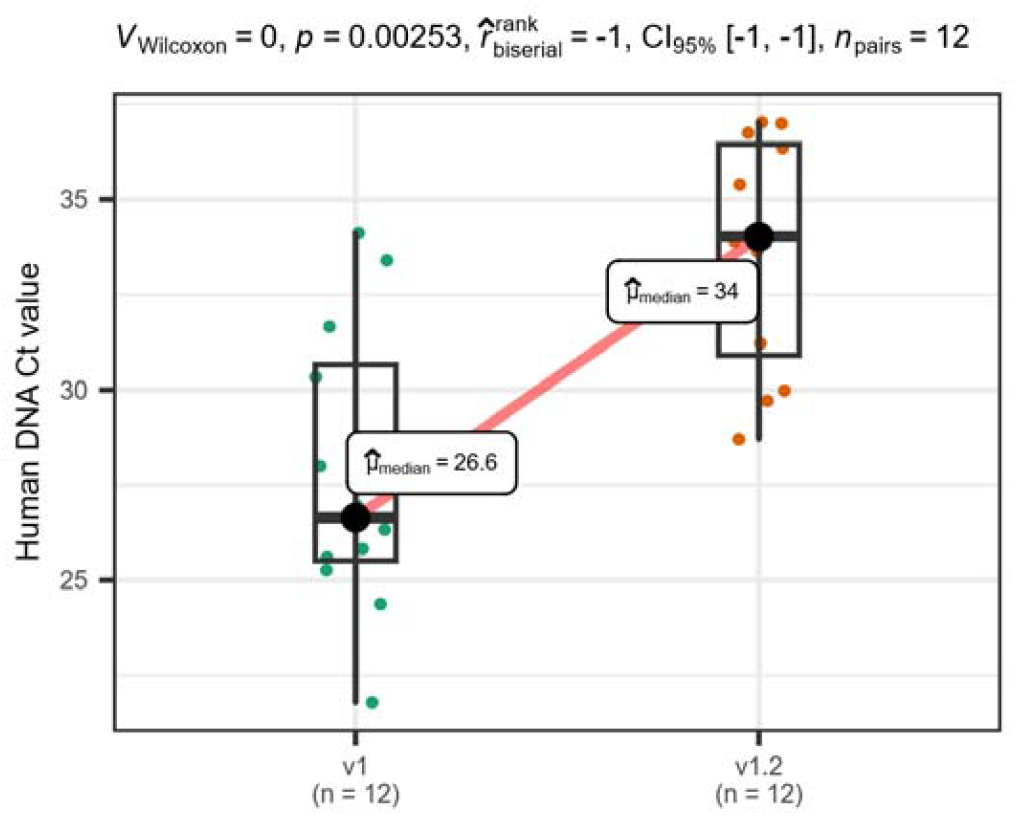
A boxplot displaying the difference between Ct values of human DNA from a qPCR targeting the RNA polA gene from a subset of 12 samples prepared by either protocol v1 or protocol v1.2. Results from a Wilcoxon signed-rank test comparing the two groups is displayed at the top of the plot.

### Organism enrichment and increased genome coverage in protocol v1.2 prepared samples

We also compared whether the reduction in human reads translated into an increase in pathogen reads. There was a statistically significant increase in micro-organism reads in v1.2 samples compared to their v1 paired sample (Fig. 3, Supplementary Figure 8 and 10). For bacterial and fungal reads, there was a median increase in organism reads of +13,920 per sample (median v1 organism read count = 3,880, median v1.2 organism read count = 17,800, *p* = 7.9 × 10^−6^) and a median increase in viral reads of +451 per sample (median v1 viral read count = 288, median v1.2 viral read count = 739, *p* = 3.2 × 10^−7^). A comparison of the same results split by sample type showed the statistically significant increase in bacterial and fungal reads in BAL (Δ = +13,400 reads, median v1 organism read count = 2,800, median v1.2 organism read count = 16,200, *p* = 0.013), ETT (Δ = +350, median v1 organism read count = 1,740, median v1.2 organism read count = 2,090, *p* = 0.009), NPA (Δ = +920, median v1 organism read count = 2,110, median v1.2 organism read count = 3,030, *p* = 0.047) and sputum samples (Δ = +84,600, median v1 organism read count = 32,400, median v1.2 organism read count = 117,000, *p* = 0.011) (Supplementary figure 9). There was also a median increase in viral reads in BAL, NPA and NTS samples but there was only a statistically significant increase in ETT samples (Δ = +6041 reads, median v1 viral read count = 499, median v1.2 viral read count = 6,540, *p* = 2.1 × 10^−6^) (Supplementary figure 11).

**Fig. 3.**
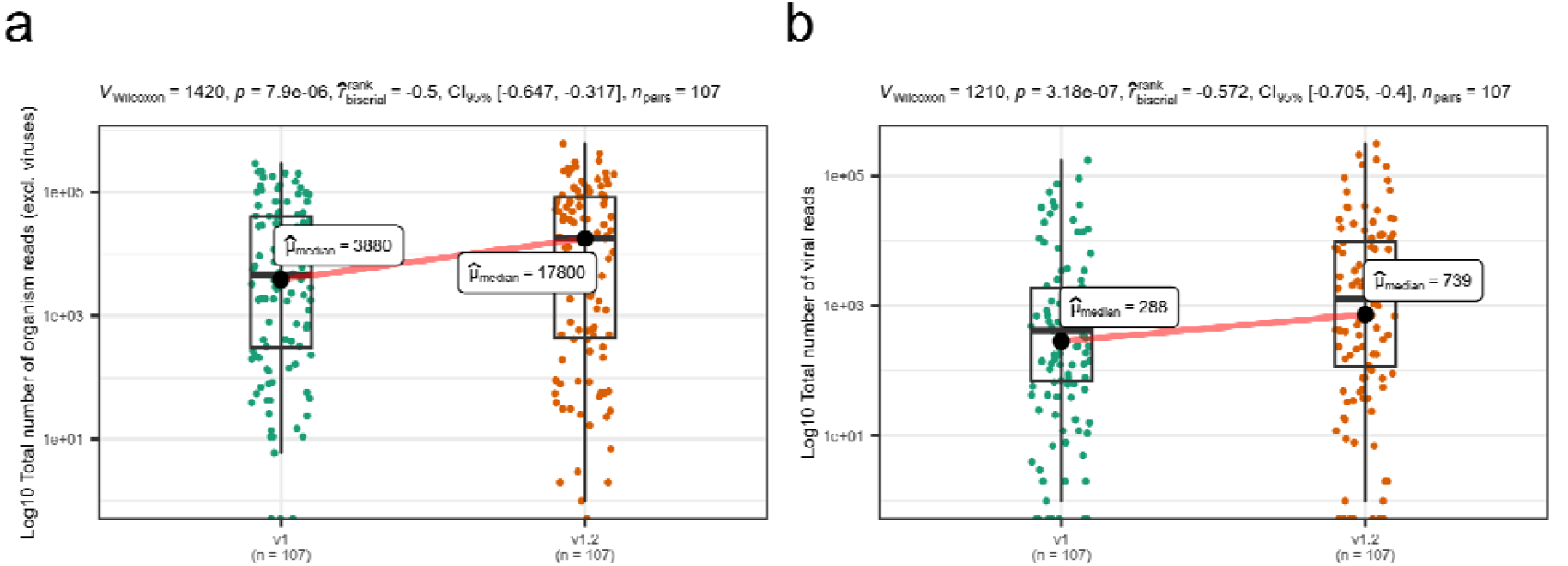
Boxplots displaying the difference between 107 samples prepared by either protocol v1 and protocol v1.2 for a) the Log10 total number of reads classified as any organism (excluding viruses). b) The log10 total number of reads classified as any virus in each sample. Results from a Wilcoxon signed-rank test comparing the two groups is displayed at the top of the plot.

To ascertain whether the increase in organism reads improved organism genome coverage and coverage read depth between protocol v1 and v1.2 samples, we computed coverage of 26 species of bacteria (Table 4), fungi and viruses present across 107 paired samples. Many of these organisms, especially viruses, were present in too few samples for meaningful comparison so results are grouped together as either bacteria (n = 5), fungi (n = 3), DNA viruses (n = 6) or RNA viruses (n = 12) for comparison and results for individual organisms can be found in Supplementary Materials (Supplementary Figures 12–15). There was an increase in genome coverage for bacteria, fungi, DNA viruses and RNA viruses even at 50X read depth for protocol v1.2 prepared samples compared to v1 (Fig. 4). At 1X read depth mean bacterial genome coverage increased from 57% in v1 samples to 69% in v1.2 and mean fungal genome coverage increased from 1% in v1 to 3% in v1.2. For DNA viruses, genome coverage increased from 55% to 62% while for RNA viruses it increased from 69% to 78%. Both increases in coverage for bacteria and RNA viruses were statistically significant (Δ = +12%, *p* = 0.0001 and Δ = +9%, *p* = 0.04, respectively by paired *t*-test). When broken down into individual organisms those that saw the greatest improvement in coverage included *Streptococcus pneumoniae* (Supplementary figure 13), RSV, Rhinovirus B and seasonal coronaviruses HCoV-OC43, HCoV-NL63 and HCoV-229E (Supplementary figure 15).

**Fig. 4.**
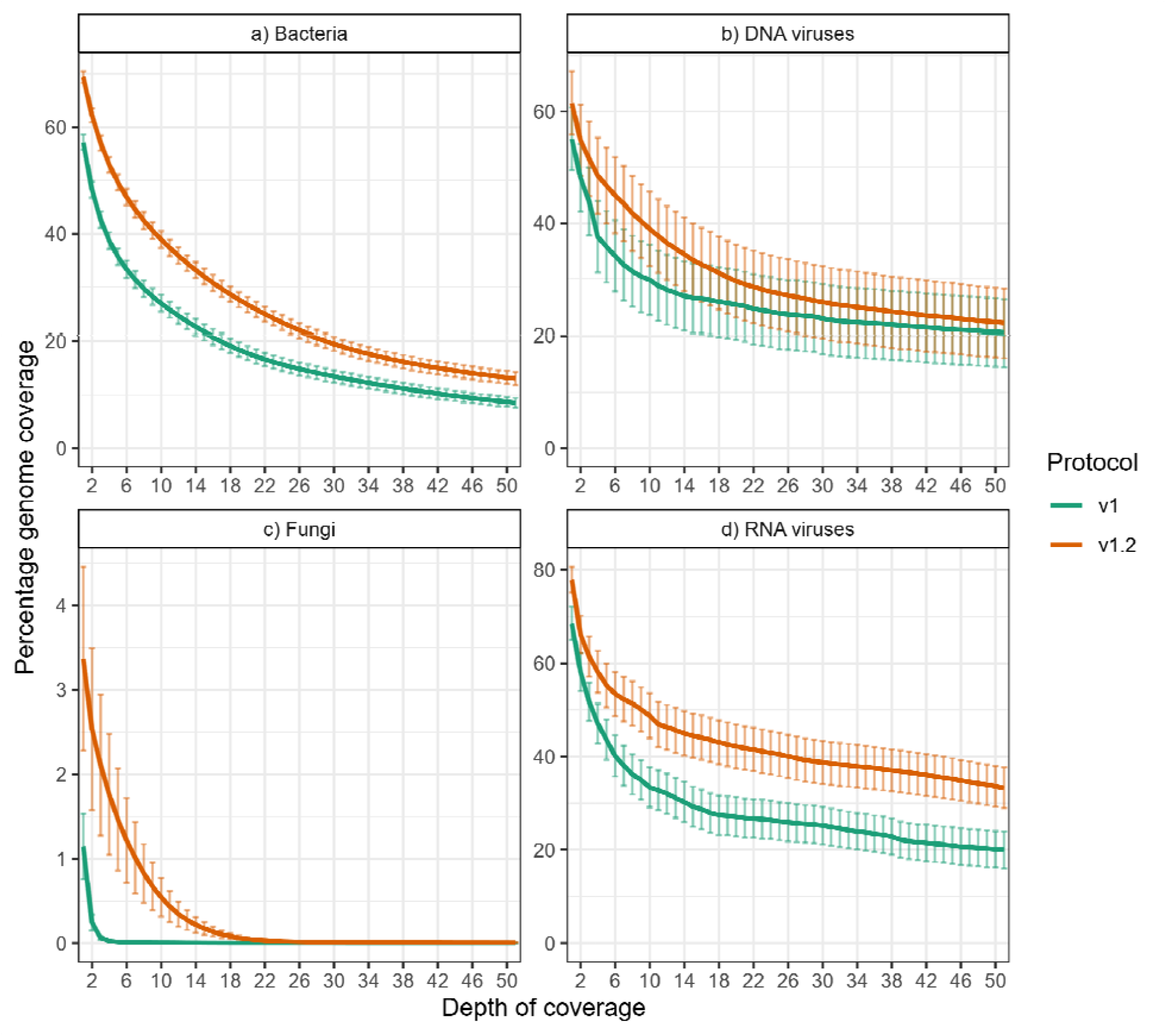
The proportion of organism genomes covered by a given read depth grouped by pathogen type and compared between protocols. The solid lines and bars represent the mean value and standard error for genome coverage at each read depth of coverage. Plots are shown for a) 5 bacteria (n = 230), b) 3 fungi (n = 86), c) 6 DNA viruses (n = 20) or any of the d) 12 RNA viruses (n = 48) included.

### Sensitivity and specificity against clinically reported organisms

We wanted to assess whether the improvements to human depletion and pathogen read recovery translate to improvements in test performance. We compared sensitivity and specificity of both v1 and v1.2 assay results to the clinically reported organisms found in each sample by orthogonal methods, as previously described^7^. We found increased sensitivity for v1.2 over v1 for bacteria and viruses whether reporting at 2 hours and 24 hours (Tables 2, and 3). At 24 hours sensitivity (95% CI) for clinically reported bacteria for v1.2 was 98% (88% – 100%) compared to 93% (81%-99%) for v1 (Table 2). This included extra detections of two *Staphylococcus aureus* including a confirmed Methicillin-resistant *Staphylococcus aureus* (MRSA) and *Pseudomonas aeruginosa* all missed by protocol v1. Viral sensitivity at 24 hours (95% CI) for clinically reported viruses was 80% (64%-91%) for v1 compared to 90% (76%-97%) for v1.2 (Table 3) and this included extra detections of SARS-CoV-2, HCoV-OC43, HPIV-2, HPIV-4 and Rhinovirus A.

**Table 2.**
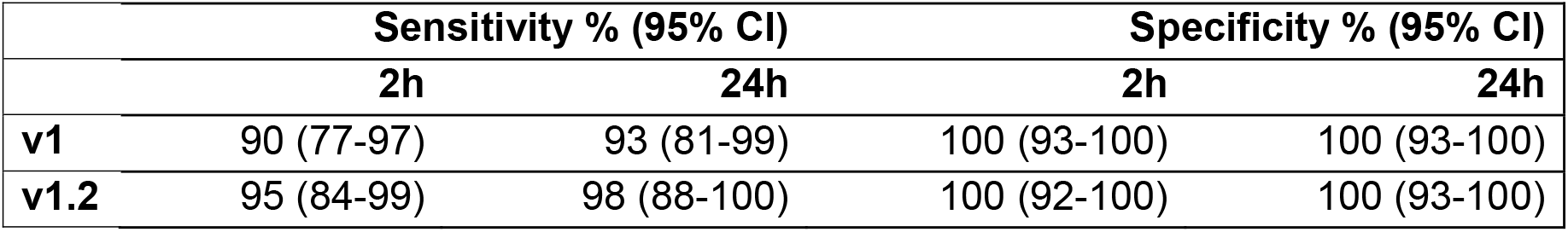
Bacterial sensitivity and specificity of metagenomics result against clinical reporting organism at 2 and 24 hours after the sequencing run has started.

|  | Sensitivity % (95% CI) |  | Specificity % (95% CI) |  |
| --- | --- | --- | --- | --- |
|  | 2h | 24h | 2h | 24h |
| **v1** | 90 (77-97) | 93 (81-99) | 100 (93-100) | 100 (93-100) |
| **v1.2** | 95 (84-99) | 98 (88-100) | 100 (92-100) | 100 (93-100) |

**Table 3.** Viral sensitivity and specificity of metagenomics result against clinical reporting organism at 2 and 24 hours after the sequencing run has started.

|  | Sensitivity % (95% CI) |  | Specificity % (95% CI) |  |
| --- | --- | --- | --- | --- |
|  | 2h | 24h | 2h | 24h |
| **v1** | 63 (46-78) | 80 (64-91) | 100 (94-100) | 100 (94-100) |
| **v1.2** | 68 (51-81) | 90 (76-97) | 98 (90-100) | 98 (90-100) |

Specificity was identical across both versions of the protocol at 2-hour and 24-hour reporting timepoints for bacteria (Tables 2). Specificity (95% CI) for viruses at 2-hour and 24-hour was 98% (90%-100%) for v1.2 compared to 100% (94%-100%) protocol v1 due to an extra detection of HSV-1 which was reported negative by PCR.

### Assay variables affecting organism recovery in protocol v1.2

A comparison of performance metrics between sites using the updated v1.2 protocol highlighted two major factors that affected the recovery of both bacterial and viral reads. Firstly, the use of different extraction platforms affected the recovery of either bacteria or viruses after sequencing of the controls (Supplementary figure 16) and secondly, the viability of samples decreased over time. We excluded samples that were processed after being refrigerated for more than 3 days after collection but we still found a 12-fold decrease in viral reads after 2 days storage at fridge temperatures and a 76-fold decrease in viral reads after 3 days (Supplementary figure 17).

## Discussion

In this multicentre validation study, we have shown significant improvements in a new version (v1.2) of our clinical metagenomics assay over the previous version (v1). Optimisation of the human depletion step and nucleic acid purification led to a lower number of human reads sequenced, higher number of pathogen reads, greater depth and coverage of pathogen genomes, and increased pathogen sensitivity. This was achieved through the deployment of the protocol across six separate hospital clinical laboratories for the first time, demonstrating this protocol’s accessibility and feasibility as a routine clinical test with a same-day turnaround time.

The initial v1 protocol was developed to enable the simultaneous detection of DNA and RNA microorganisms in a single reaction tube. This was in contrast to other common metagenomic assays that require users to split samples for separate DNA and RNA analyses^8,9^. In the validation of the v1 protocol against 50 clinical samples at a single site, we demonstrated 94% sensitivity and 100% specificity for the detection of viruses and bacteria^6^. During clinical implementation at the same site using external quality controls, the v1 protocol maintained comparable performance achieving 97% sensitivity and 98% specificity for bacterial detection, 89% sensitivity and 100% specificity for viral detection, and 89% sensitivity and 100% specificity for fungal detection^7^.

Despite these promising results, the validation study with protocol v1 showed limitations in genome recovery, with only 42% of viruses and 15% of bacteria yielding sufficient genome coverage for downstream analysis. Although the protocol achieved a substantial 256-fold reduction in human DNA background, it was hypothesised that further improvements, particularly in human DNA depletion, could enhance overall performance, especially in terms of viral detection and microbial genome recovery.

To enhance human DNA depletion, we first modified the protocol to bead-beat the sample twice, which we hypothesised would improve lysis efficiency of human cells and thereby increase the amount of accessible host DNA available for enzymatic digestion.

Next, we identified that a weakness in the original protocol was the endonuclease enzyme used for host DNA digestion. The HL-SAN enzyme requires a high-salt buffer to achieve optimal activity, conditions previously validated for the detection of DNA microorganisms^10^. However, this buffer was found, during the development of the protocol, to compromise the integrity of enveloped RNA viruses, leading to reduced detection efficiency and therefore was not used in the v1 of the protocol. To improve performance, HL-SAN was replaced with M-SAN, a modified endonuclease engineered to function optimally without requiring the high-salt buffer^11^. In addition, a critical refinement was introduced: the M-SAN reaction was deliberately stopped prior to the extraction step. This interruption prevented concurrent digestion of microbial DNA/RNA during extraction, due to HLSA activity which is still active at ambient temperature expecting HLSAN to be active after the incubation step, an issue not previously addressed in our workflow, and contributed to the preservation of microbial genomes. Collectively, these changes have directly led to a 7% increase in sensitivity in detecting bacteria and 10% increase in sensitivity in detecting viruses at 24 hours reporting. In addition to this we are now recovering a larger proportion of bacterial, fungal and viral genomes than previously (Fig. 4), and detecting RNA viruses at Ct 35, improving the possibilities of additional pathogen typing and validating antimicrobial resistance profiling.

The improved protocol (v1.2) was validated across six different hospitals using three distinct nucleic acid extraction methods, with a total of 107 clinical samples spanning various respiratory specimen types. This multi-site validation shows the robustness, adaptability, and reproducibility of the method in real-world clinical settings even using different extraction platforms, which can substantially influence the number of reads generated downstream (supplementary figure 16). Validation with over 100 samples enabled a meaningful assessment of diagnostic performance and confirmed the assay’s reliability across workflows and institutions.

Importantly, this method also enabled the detection of a parasite, *Toxoplasma gondii*, for the first time with increased number of reads in v1.2 of the protocol (Patient number 100, sample number 200, supplementary table), highlighting the true agnostic nature of this approach. Unlike targeted assays, which require prior assumptions about likely pathogens, this unbiased metagenomic strategy offers comprehensive detection, making it especially useful in cases of uncommon or unexpected infections.

Limitations of this protocol remain the requirement to process fresh rather than frozen samples to prevent additional microbial loss during thawing and the use of samples not older than 48h (Supplementary figure 17. The potential for microorganisms present in low relative abundance in polymicrobial samples to go undetected due to the competitive nature of sequencing, and the inability to capture free nucleic acid that qPCR can detect, are issues that have been consistently reported by other groups using various depletion and sequencing methods, highlighting them as inherent constraints of the technique ^12–15^. This workflow was originally developed and validated using Oxford Nanopore Technologies (ONT) R9 chemistry, as the ONT Q-LINE sequencing platforms for clinical laboratories do not yet permit sequencing with R10 flow cells. We have subsequently confirmed this assays compatibility with R10 chemistry and future work will migrate the protocol with the ONT release of the next versions of the Q-LINE.

Beyond broad pathogen coverage and the limitations, this single-tube protocol offers short turnaround times, with results available in six and seven hours from sample receipt for detection of microbial DNA and RNA. This makes the method suitable for both urgent clinical decision-making and infection control, facilitating timely patient management and public health interventions. Further improvements to the method brings metagenomics closer as a replacement for targeted PCR and culture.

## Online Methods

### Version 1.0 and version 1.2 protocols and differences

Both version 1.0 and 1.2 protocols can be found in their entirety on protocols.io at https://dx.doi.org/10.17504/protocols.io.eq2lyw5qpvx9/v2. For comparisons made in this paper each sample was either processed using the original protocol (v1.0), with modifications described^16^, including the introduction of quality controls. The second aliquot was processed using the updated protocol (v1.2). The key differences in v1.2 include: (1) a double bead-beating step to enhance lysis of human cells, and (2) the replacement of HLSAN with MSAN, supplemented with Mg^2^? to optimise enzyme activity. The reaction was then stopped with EDTA, following the manufacturer’s instructions. All other steps in the protocol remained unchanged.

### Human DNA depletion assessment on control material

To evaluate the impact of human DNA depletion on the detection of viral and atypical bacterial targets, Respiratory Panel 2.1 (RP2.1) Controls (Zeptometrix, 12 x 0.3mL), Control 1 and Csontrol 2, were pooled to obtain a final volume of 1,500 µl. The combined sample was thoroughly vortexed and divided into three aliquots. Two 500 µL aliquots were subjected to the human DNA depletion protocols: one using version 1 (v1) and the other using version 1.2 (v1.2). The remaining 200 µl aliquot was directly extracted without any depletion step using the MagNA Pure system (Roche®).

### Human DNA depletion assessment on clinical samples

For the validation and comparison of protocol versions 1.0 and 1.2, a total of 107 surplus clinical samples were processed in parallel. Collection of surplus samples and linked clinical data was approved by South Central-Hampshire B REC (20/SC/0310). The sample types included bronchoalveolar lavage (BAL, n=40), endotracheal tube aspirates (ETT, n=37), nasopharyngeal aspirates (NPA, n=6), nasal/throat swabs (NTS, n=7), pleural fluids (PFL, n=2) and sputum (SPT, n=15). These samples were collected from six different hospitals: Site 1 (n=11), site 2 (n=27), site 3 (n=34), site 4 (n=12), site 5 (n=11) and site 6 (n=12).

Sites 1, 2 and 5 used the EZ1 Advanced XL platform. The extraction was performed extracting 200 µL of sample using the Qiagen EZ1 Virus Mini Kit v2.0, with a final elution volume of 60 µL. For site 1, extraction was performed comprising 200 µL of sample mixed with 200 µL of ATL buffer. In all the laboratories the carrier RNA was replaced with AVE buffer during the process.

Site 3 used MagNA Pure 24 Instrument (Roche). In a purification (cartridge) run, the reagent cassette used was Total NA isolation kit 1.1 for the Bronchoalveolar lavage sample type. The input volume selected was 200 μl, and the extraction protocol was Fastpathogen 200 1.1.

Site 4 used the KingFisher™ Apex instrument with MagMAX™ Viral/Pathogen II (MVP II) Nucleic Acid Isolation Kit. Samples were mixed with lysis buffer and incubated at 20°C for 15 minutes before adding Proteinase K and binding beads. Wash, elution, and tip comb plates are prepared separately: wash buffer, 80% ethanol, and elution buffer.

Site 6 used the QIAamp UCP Pathogen Mini kit. The kit was used according to manufacturer’s instructions with a volume of 600 μl of lysate from each sample was transferred into QIAamp UCP Mini spin columns and eluted into 35 μl of elution buffer. The columns were left to incubate at room temperature for 10 minutes. After incubation, the columns were centrifuged at full speed (18,000 × g) for 2 minutes. This step was repeated by adding an additional 30 μl of elution buffer and centrifuging again.

Samples were only included in the analyses if they were processed <3 days after collection.

### Human RNA *polA* gene quantitative PCR

To assess the effectiveness of human DNA depletion, a subset of 27 respiratory samples from the 107 collected, including 13 bronchoalveolar lavage (BAL), 9 sputum, 2 endotracheal tube (ETT) aspirates, 2 nasal/throat swabs (NTS), and 1 pleural fluid (PF) were each split into two aliquots. One aliquot from each sample underwent human DNA depletion prior to nucleic acid extraction, while the other was extracted without depletion. A PCR targeting human DNA was then performed on both aliquots to compare the level of human DNA present.

### Pathogen PCR for the RP2.1 Control

Quantitative PCR was performed on the RP2.1 Control (Zeptometrix) using the VIASURE Respiratory Panel III Real-Time PCR Detection Kit (Certest Biotec) according to the manufacturer’s instructions. Each reaction was prepared in a final volume of 20 µl, including 5 µl of nucleic acid template. Thermal cycling was carried out on a QuantStudio 7 Real-Time PCR System (Thermo Fisher Scientific)

### Sample stability

To investigate the impact of storage temperature on sequencing signal, a stability experiment was conducted in which a mock sample was prepared and stored under two different temperature conditions: room temperature and refrigeration (4°C). The objective was to determine whether the independent variable, storage temperature, affected the sequencing output.

The mock sample was composed of a mixture of clinically relevant pathogens, including *Streptococcus pneumoniae* NCTC 12977, *Candida albicans* ATCC 14053, *Enterococcus faecalis, Staphylococcus aureus* NCTC 12493, *Escherichia coli, Pseudomonas aeruginosa, Mycobacterium abscessus*, and Human Herpes Virus Type 6 Stock (Quantitative) (Zeptometrix, 1 mL). Each organism was cultured from an organism-specific purity plate and adjusted to a 0.5 McFarland suspension using a densitometer. From these suspensions, defined volumes were added to the sample to achieve target concentrations representative of clinically detectable levels.

Specifically, 1 mL of *S. pneumoniae* NCTC 12977 (~1.5 × 10^8^ CFU/mL) was added to yield a final concentration of 2.5 × 10^7^ CFU/mL in the mixture. Similarly, 0.4 mL of *C. albicans* ATCC 14053 (~1.4 × 10^6^ CFU/ml) resulted in a final concentration of approximately 9.3 × 10^4^ CFU/ml. For *E. faecalis*, 0.4 ml of a ~1.5 × 10^8^ CFU/ml suspension was added to achieve 1.0 × 10^7^ CFU/ml. Lower volumes (0.004 ml each) of *S. aureus* NCTC 12493, *E. coli, P. aeruginosa*, and *M. abscessus*, all adjusted to ~1.5 × 10^8^ CFU/ml, were included to yield final concentrations of approximately 1.0 × 10□ CFU/ml for each. Additionally, 0.3 ml of herpesvirus stock (1.0 × 10^6^ copies/ml) was included, resulting in a final viral load of 5.0 × 10^4^ copies/ml.

The total volume of the mock sample was brought to 6 mL by combining 2,116 µL of the organism suspension mix with 3,884 µL of microbial DNA-free water, which served as the negative control matrix. This final mixture was then split into two equal aliquots of 3 mL each. One aliquot was stored at room temperature, while the other was refrigerated. Both were processed at defined time points to assess the stability of the sequencing signal under the two storage conditions.

### Comparison of the extraction platform using RP2.1 Control

An aliquot of 900 µL was prepared by mixing *Control 1* and *Control 2* from the RP2.1 Control. This was used to compare four extraction instruments: EZ2™ (Qiagen®), KingFisher™ (Thermo Fisher Scientific™), MagNA Pure 24™ (Roche®), and V-Quick™ (Certest Biotec®). Each instrument was paired with its corresponding extraction kit: Virus Mini Kit v2.0 (Qiagen®), MagMAX™ Viral/Pathogen II (MVPII) Nucleic Acid Isolation Kit (Thermo Fisher Scientific™), MagNA Pure 24 Total NA Isolation Kit (Roche®), and Viasure® Universal Pathogen Extraction Kit (Certest Biotec®).

Following extraction, the v1.2 protocol was applied, and the samples were sequenced on the same flow cell to eliminate variability between flow cells. The procedure was repeated three times, each time using different barcodes to minimise barcode-associated variability.

### Sequencing and bioinformatic analysis

Sequencing was performed on 9.4.1 flow cells on GridION Q (Oxford Nanopore Technologies, Oxford, UK). Sequencing data was basecalled and demultiplexed using the ‘barcode at both ends’ setting using guppy (version 6.1.5) within MinKNOW (version 22.05.7). Reads with a Q score below 10 were filtered. Data were analyzed at two hours and 24 hours using a previously described bioinformatics pipeline^7^. The pipeline can be found at https://github.com/GSTT-CIDR/metagenomics_workflow/releases/tag/v1.0.

Reference genomes for a curated panel of 26 clinically relevant organisms (Table 4) were downloaded from NCBI RefSeq using the datasets tool (v15.10.0) with the -- reference option to ensure high-quality assemblies.

**Table 4.**
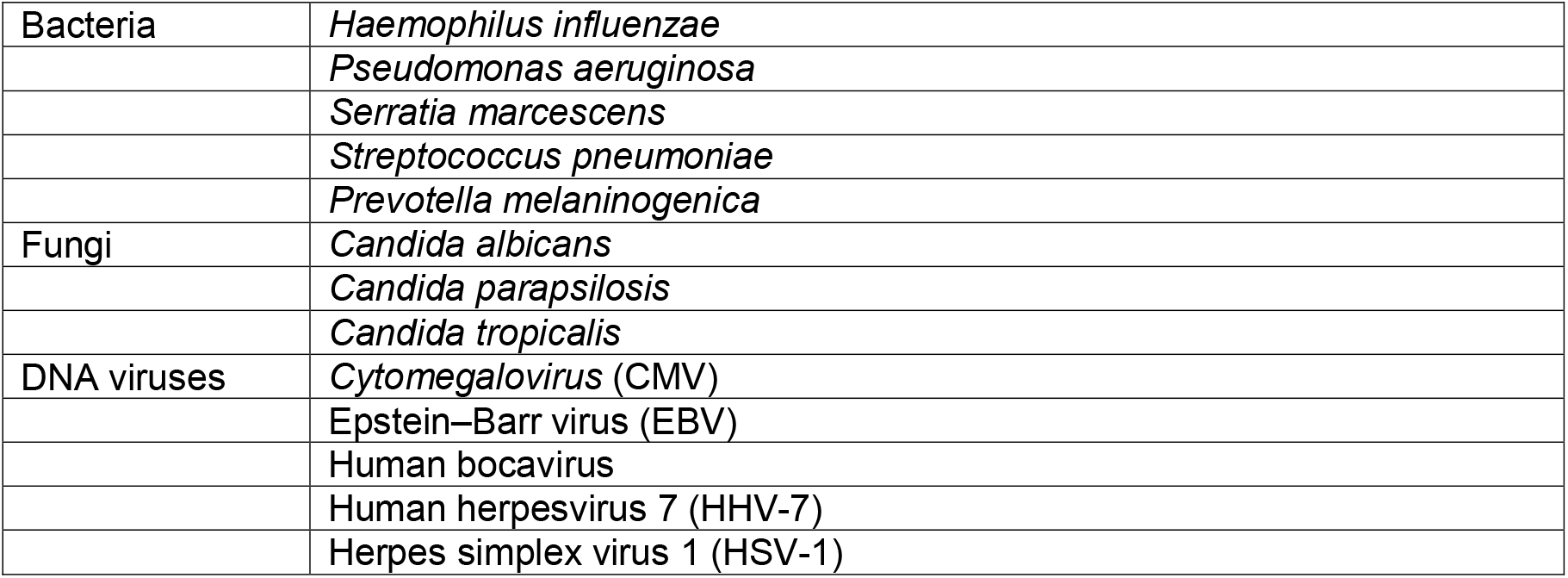
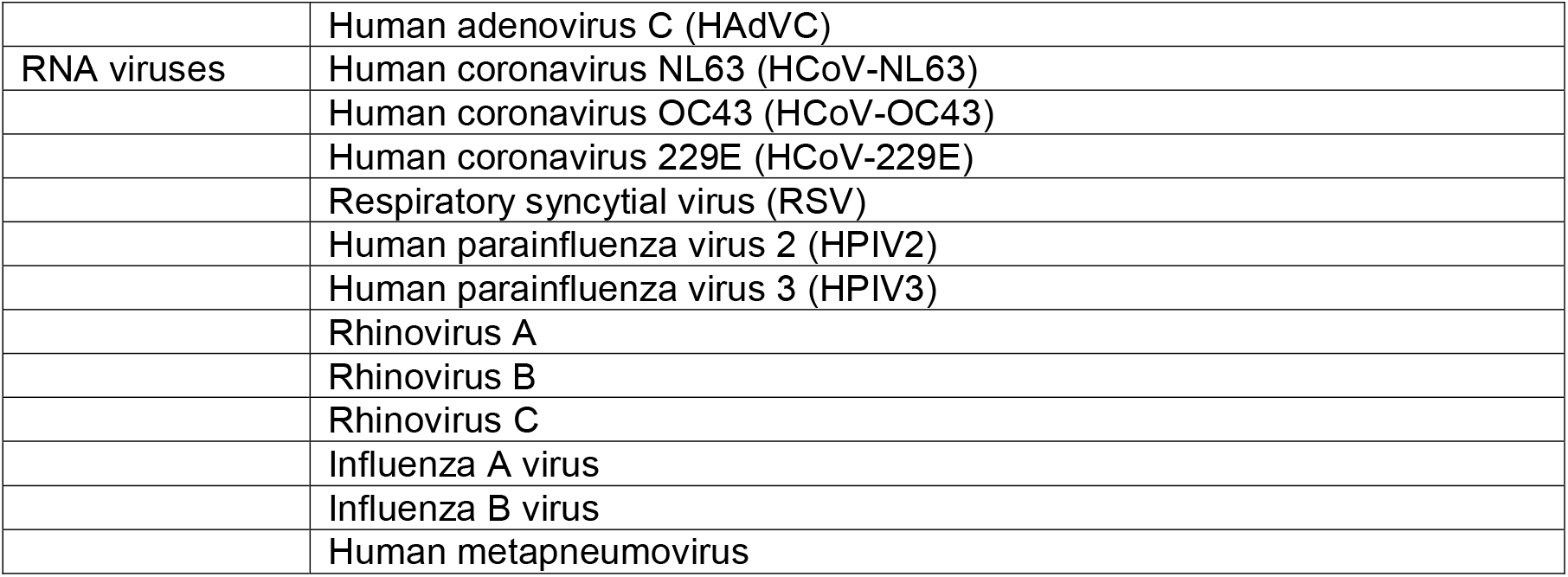
List of 26 organisms included in reference based mapping comparison of genome coverage.

Organism-specific read mapping was performed for each taxon using Minimap2^17^ (v2.24). Only reads belonging to samples identified as positive for the target organism by the metagenomics pipeline were mapped. Qualimap2^18^ (v2.2.1) was used to generate genome coverage statistics for each mapped BAM file.

### Statistical analysis

Based on prevalence from our previous study^7^, expecting a sensitivity of 0.9, specificity of 0.95, failure rate of 5%, and prevalence of bacterial detections at 50% a sample number of ≥70 was pre-specified for adequate power with ±10% precision in performance calculations (https://wnarifin.github.io/ssc/sssnsp.html). For viruses, based on a previous prevalence of detections at 35% a sample number of ≥99 was needed for adequate power with ±10% precision in performance calculations. Power calculations precluded fungi from being included in performance calculations due to low prevalence of detections at 5%. All statistical analyses were carried out using R (v4.4.3) and data was processed using tidyverse (v2.0.0) and ggstatsplot (v0.13.0). Paired comparisons between v1 and v1.2 were conducted using ggstatsplot’s ggwithinstats (Wilcoxon signed-rank tests) and visualized using ggplot2 (v3.5.1). *P* values < 0.05 were considered statistically significant and 95% confidence intervals are given where appropriate. Sensitivity, specificity and positive/negative predictive values were calculated using epiR (v2.0.84) in R.

## Data Availability

All sequencing data used in this study is available from the Sequence Read archive under study accession number PRJNA1348125. Bioinformatics pipleine used for analysis is available at https://github.com/GSTT-CIDR/metagenomics_workflow/releases/tag/v1.0

https://www.ncbi.nlm.nih.gov/sra/PRJNA1348125

https://github.com/GSTT-CIDR/metagenomics_workflow/releases/tag/v1.0

## Data availability

All sequencing data used in this study is available from the Sequence Read Archive under study accession number PRJNA1348125.

## Competing Interests

JE is employed part time by Oxford Nanopore Technologies as Vice President of Medical Affairs. AAM and RB have a patent pending for the mechanical human DNA depletion method (PCT/GB2023/051417; appendix 1 p 4). All other authors declare no competing interests.

## Acknowledgements

RMD, AAM, LBS and GN were supported by Synnovis’ Innovation Accelerator Fund: Transformative Award 2023 [T004]. LBS and GN were funded by the Medical Research Council (MR/W025140/1; MR/T005416/1).

## Notes

### Competing Interest Statement

Jonathan Edgeworth is employed part time by Oxford Nanopore Technologies as Vice President of Medical Affairs. Adela Alcolea-Medina and Rahul Batra have a patent pending for the mechanical human DNA depletion method (PCT/GB2023/051417; appendix 1 p 4). All other authors declare no competing interests.

### Clinical Protocols

https://www.protocols.io/view/pan-microbial-metagenomics-protocol-v1-2-eq2lyw5qpvx9/v2

### Funding Statement

Ronan Doyle, Adela Alcolea-Medina, Luke Blagdon Snell and Gaia Nebbia were supported by Synnovis Innovation Accelerator Fund: Transformative Award 2023 [T004]. Luke Blagdon Snell and Gaia Nebbia were funded by the Medical Research Council (MR/W025140/1; MR/T005416/1).

### Author Declarations

Collection of surplus samples and linked clinical data was approved by South Central-Hampshire B REC (20/SC/0310).

### Summary of Updates

Author added and author affiliations updated. Sequence Read Archive accession number added.

