## Supplementary figures for "Optimisation of whole cell human depletion provides increased sensitivity and microbial genome coverage from respiratory metagenomic assays"

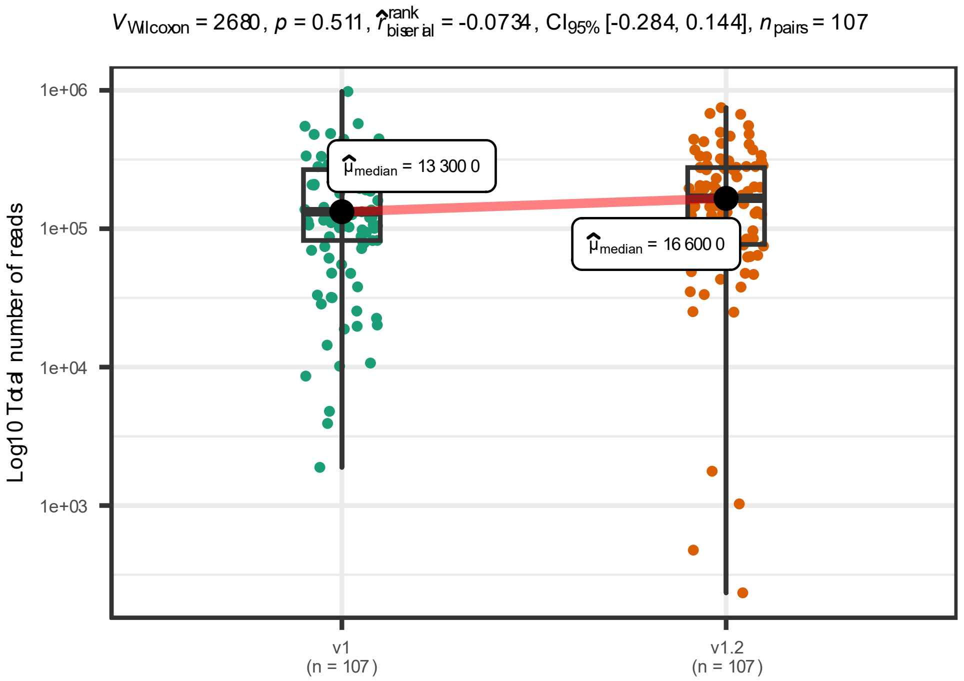

Supplementary figure 1. Total number of reads per sample generated by protocol v1 and v1.2.

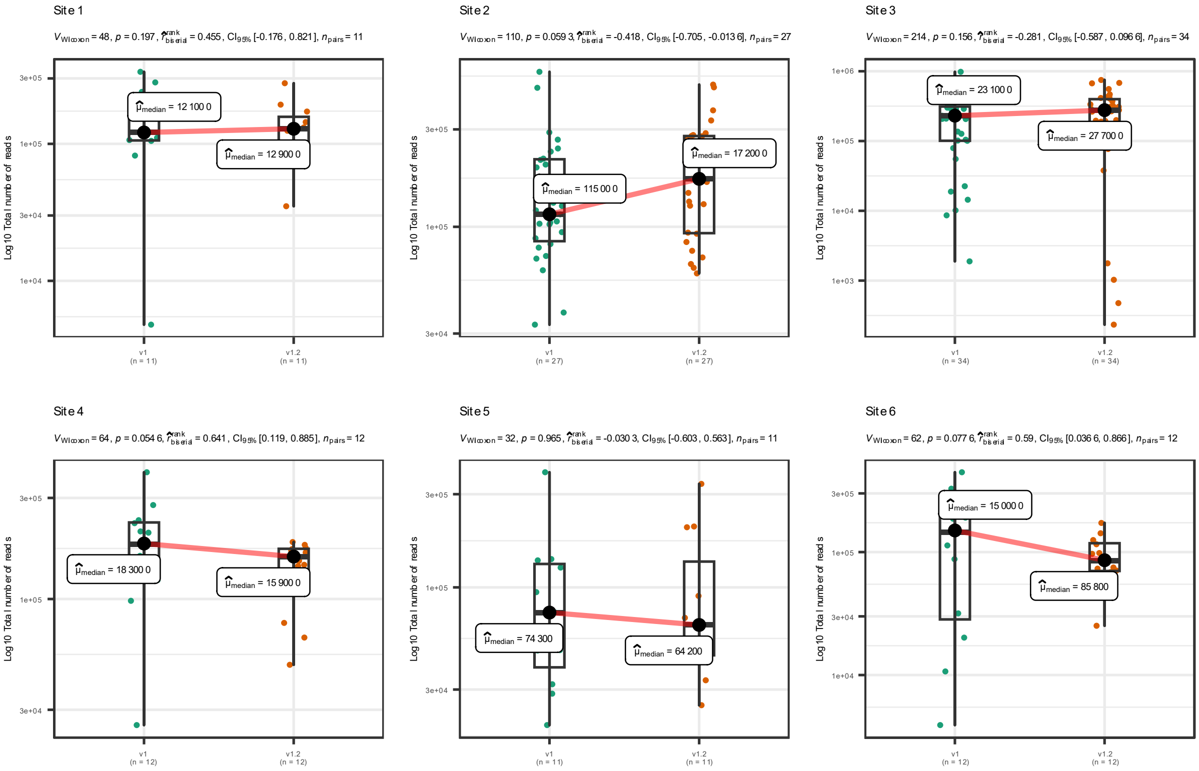

Supplementary figure 2. Total number of reads per sample by laboratory site

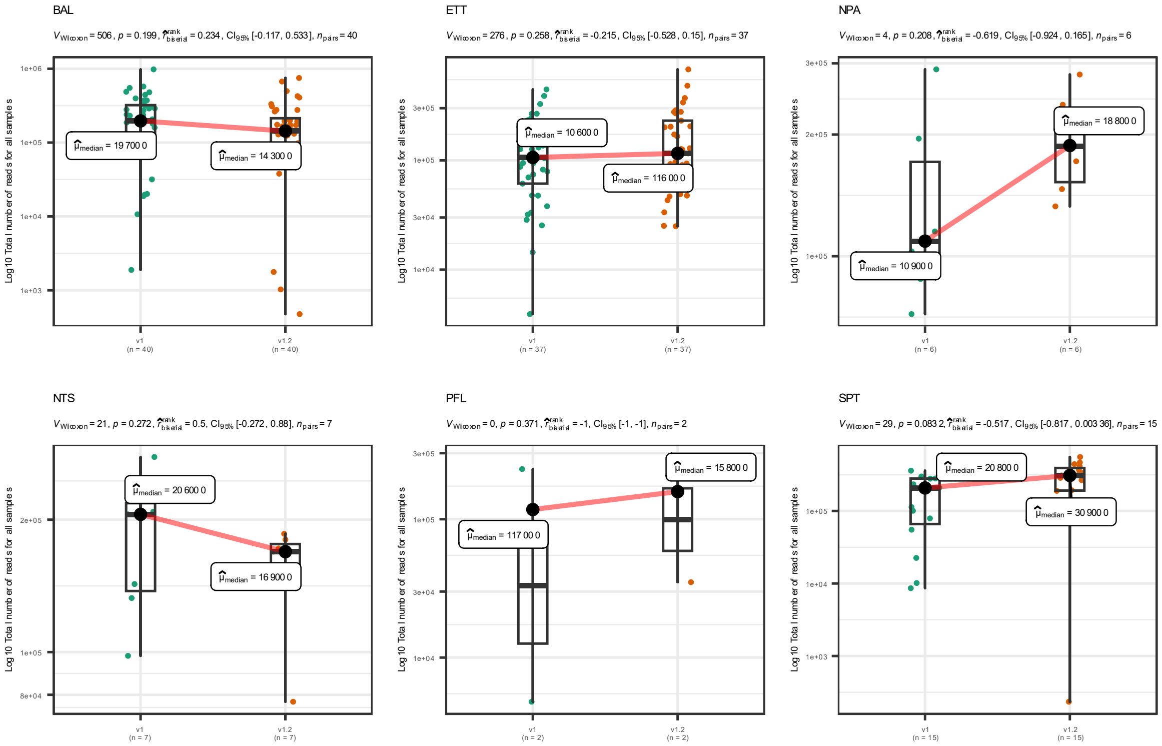

Supplementary figure 3. Total number of reads per sample shown by sample type.

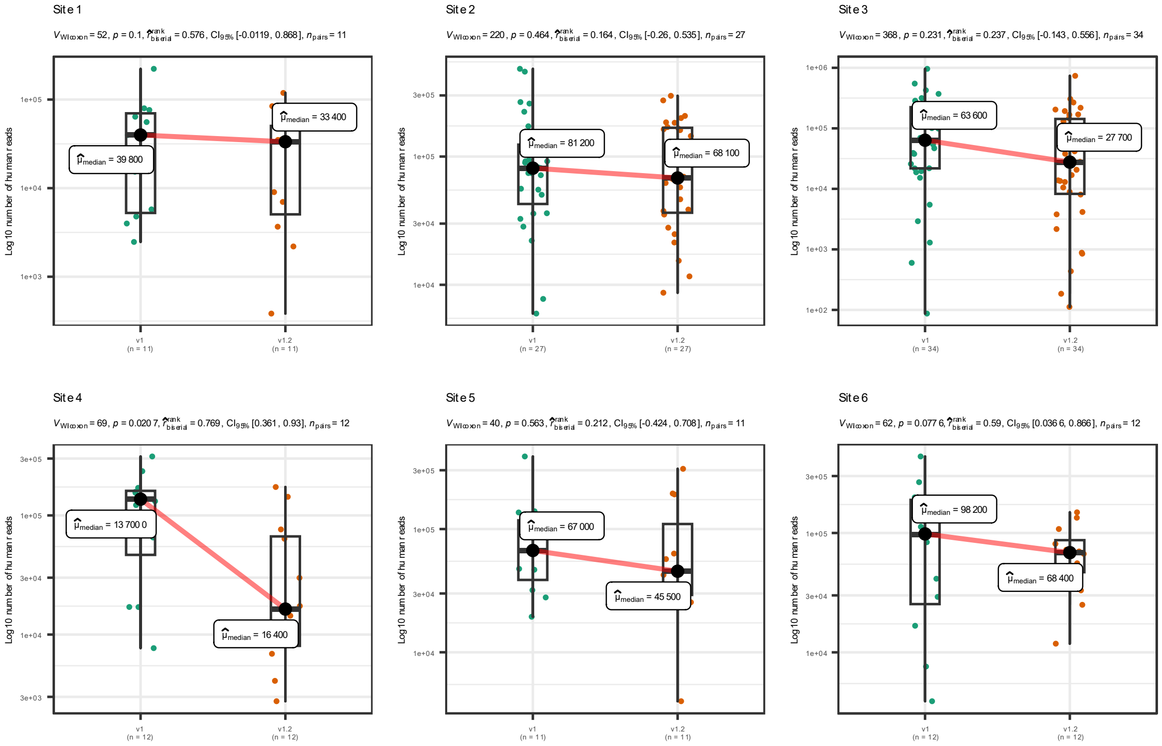

Supplementary figure 4. Total number of reads classified as human per sample by laboratory site.

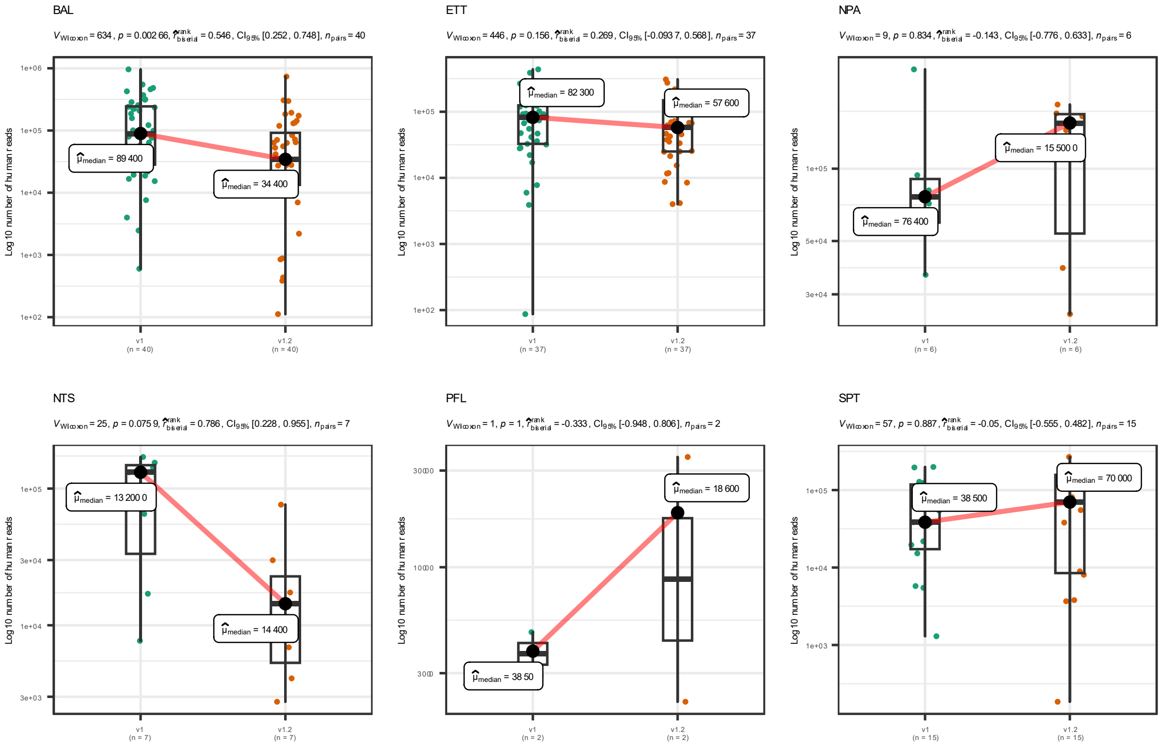

Supplementary figure 5. Total number of reads classified as human per sample shown by sample type

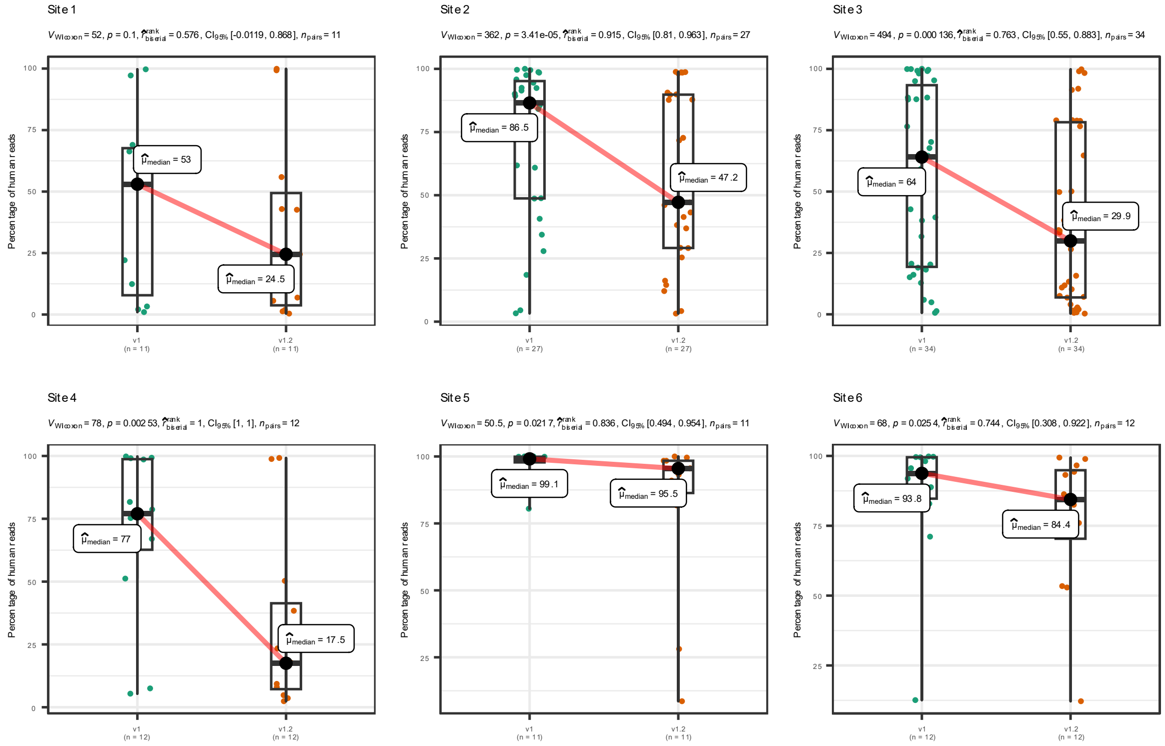

Supplementary figure 6. Percentage of human reads per sample by laboratory site

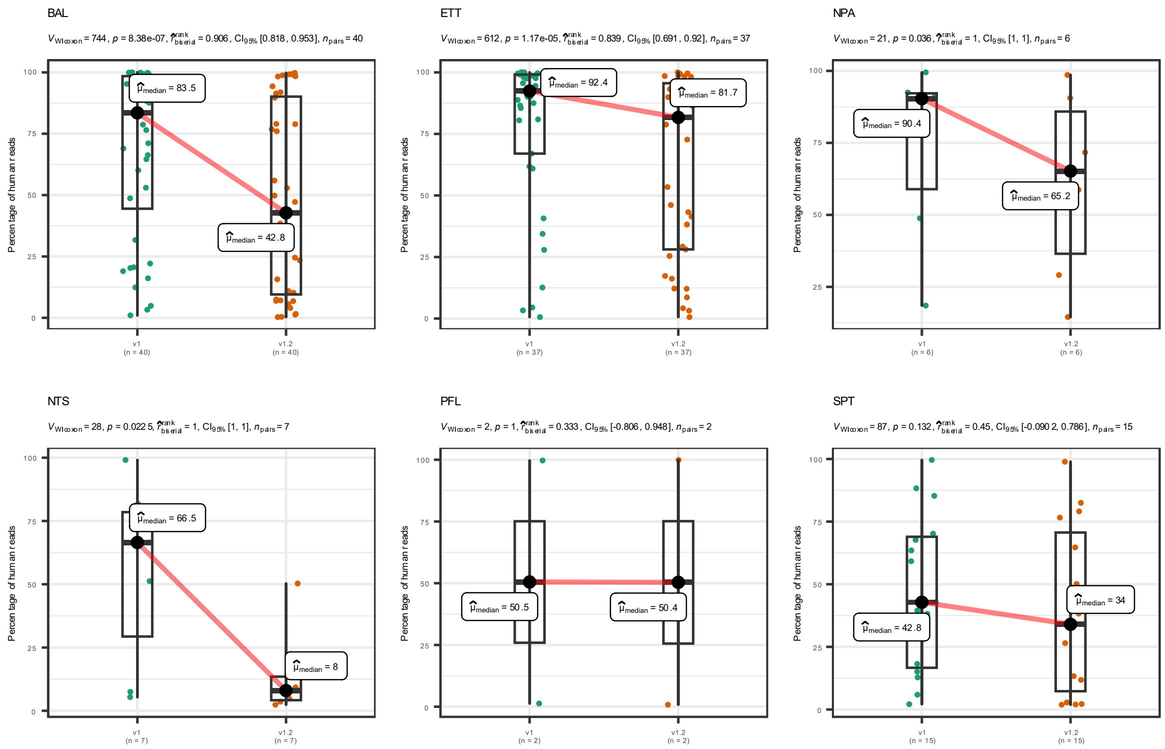

Supplementary figure 7. Percentage of human reads per sample by sample type

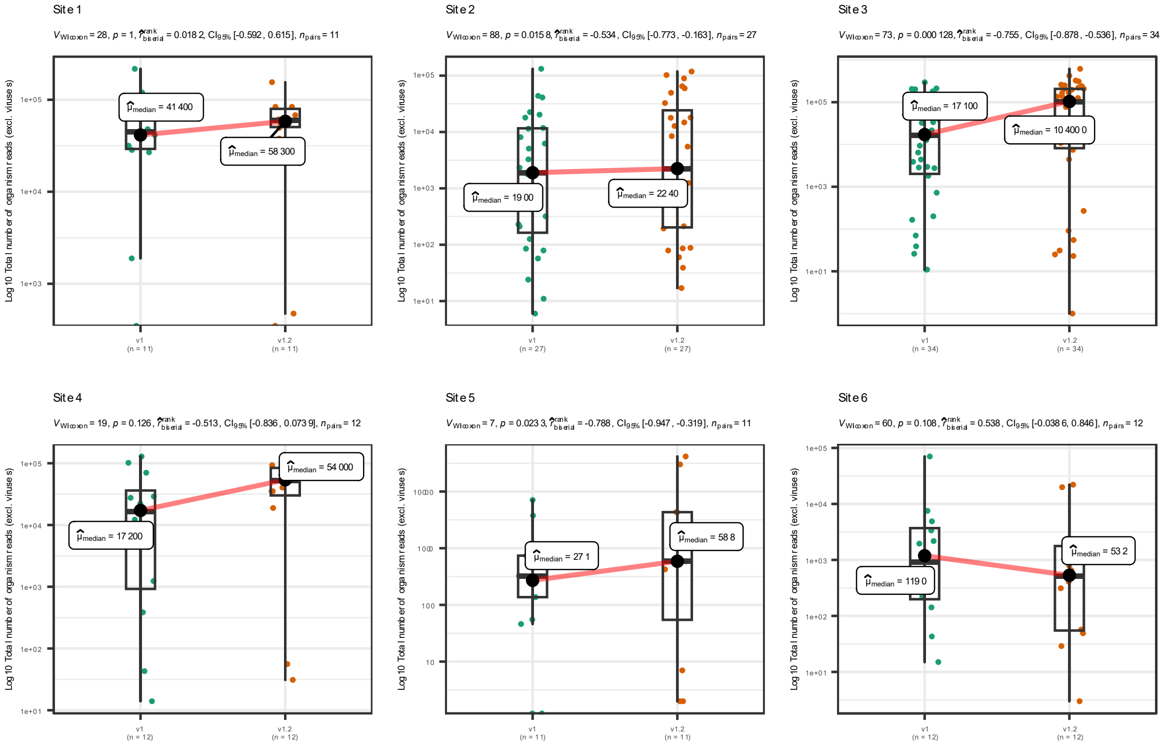

Supplementary figure 8. Total number of organism reads per sample (excl. viruses) by laboratory site.

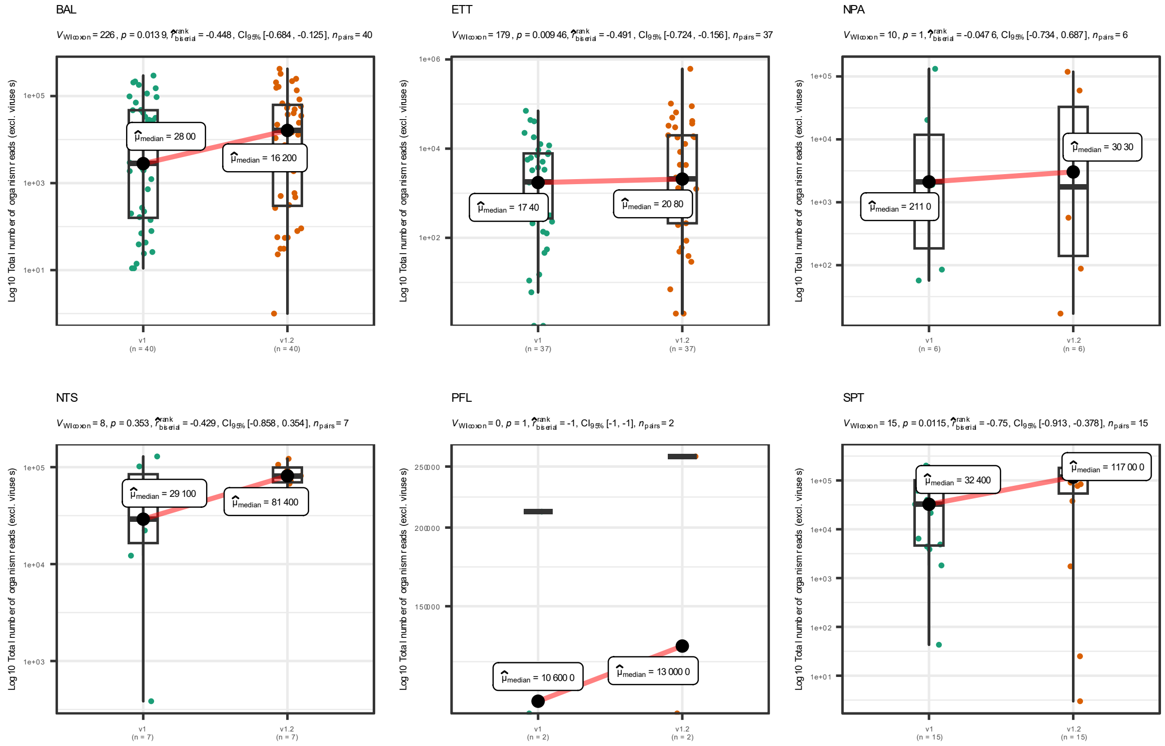

Supplementary figure 9. Total number of organism reads per sample (excl. viruses) by sample type

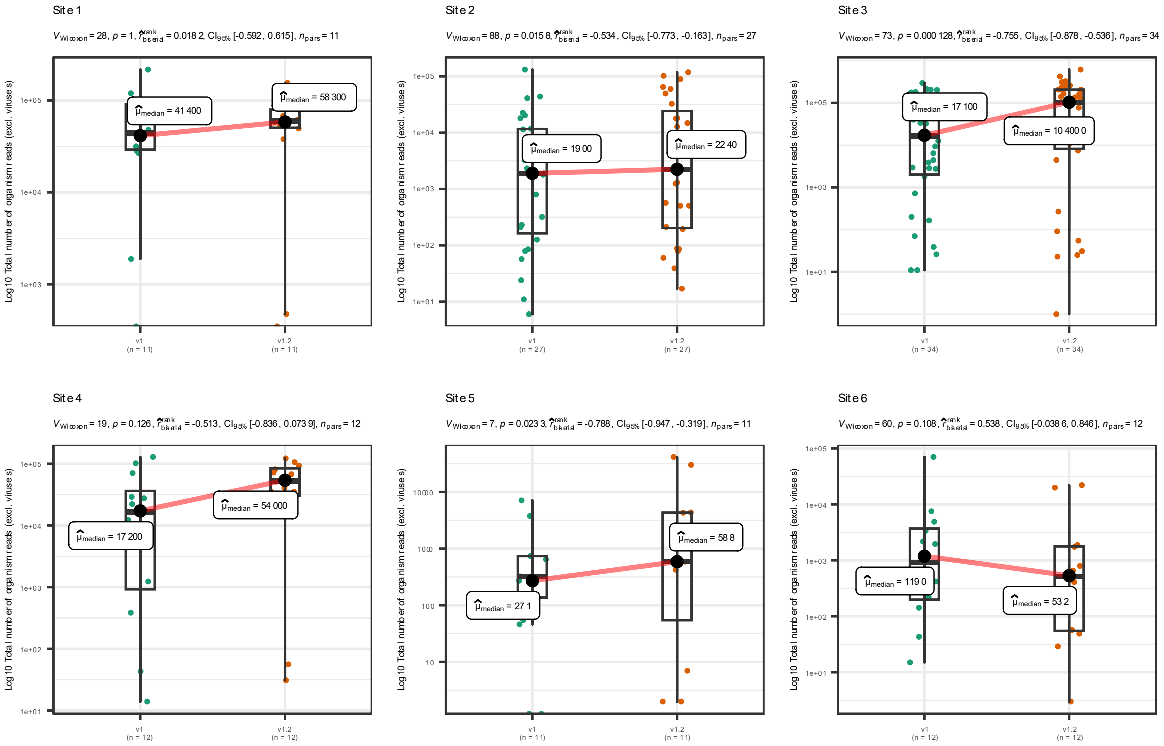

Supplementary figure 10. Total number of viral reads per sample by laboratory site.

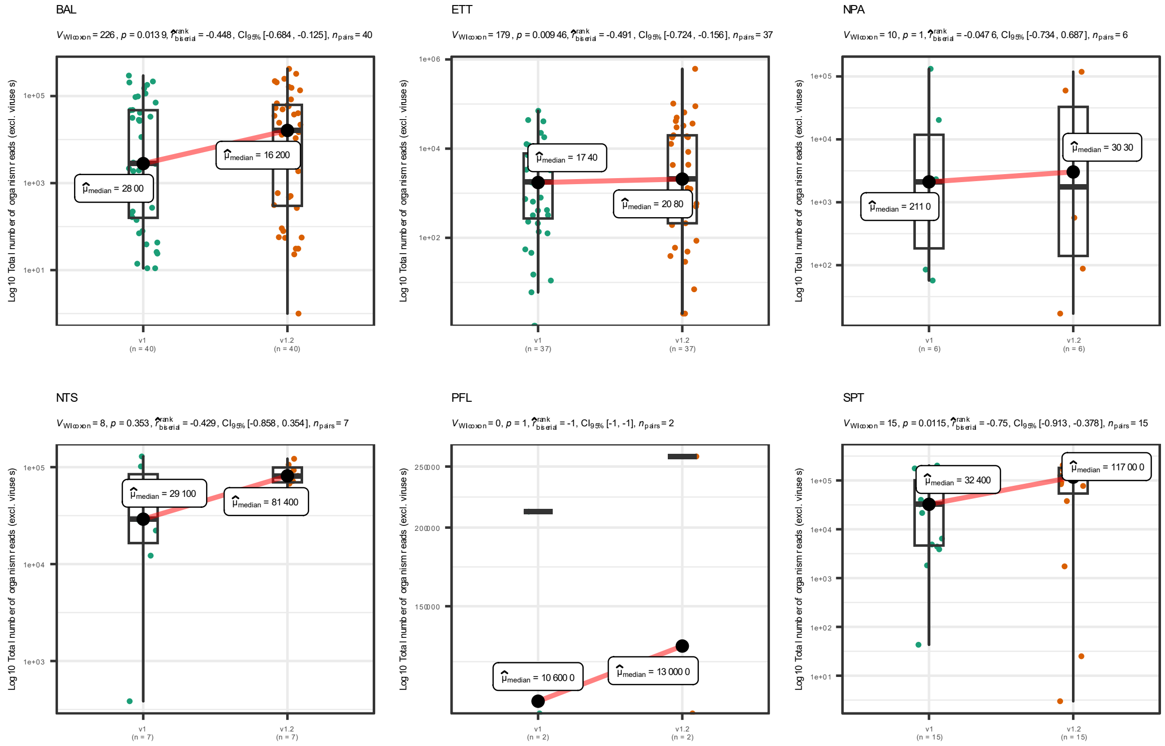

Supplementary figure 11. Total number of viral reads per sample by sample type

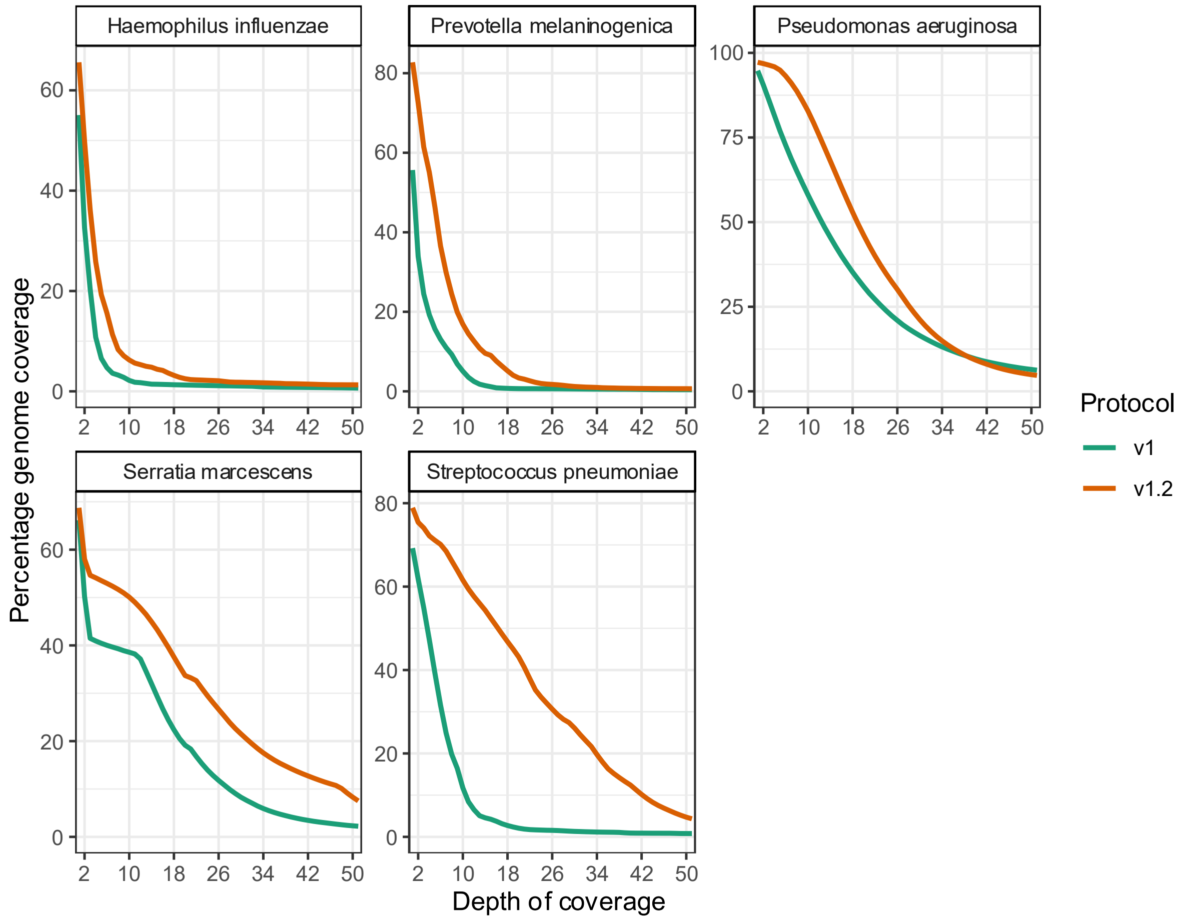

Supplementary figure 12. Median v1 and v1.2 bacterial genome coverage for individual bacteria.

Number of paired samples compared for each organism: *Haemophilus influenzae* (n = 30), *Prevotella melaninogenica* (n = 28), *Pseudomonas aeruginosa* (n = 4), *Serratia marcescens* (n = 14) and *Streptococcus pneumoniae* (n = 36).

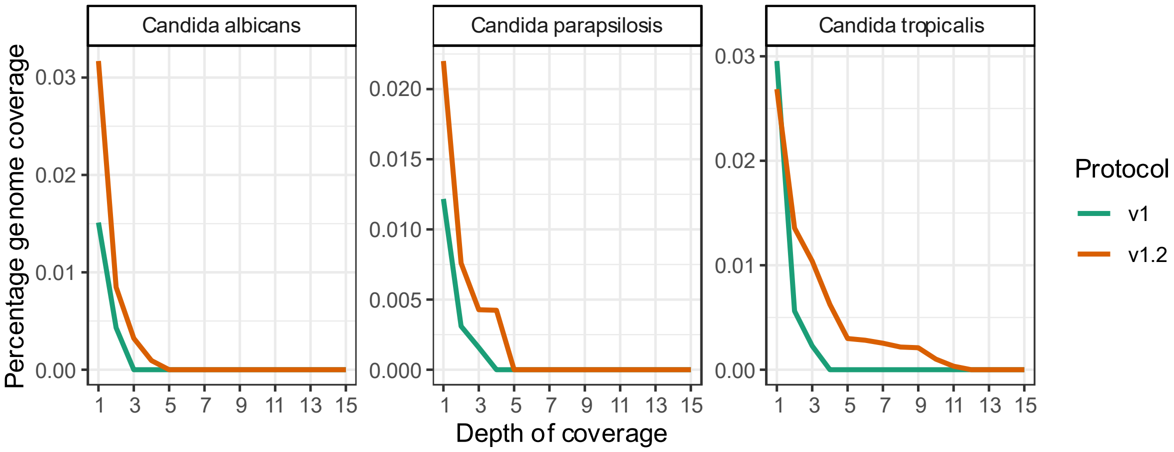

Supplementary figure 13. Median v1 and v1.2 fungal genome coverage for individual fungi.

Number of paired samples compared for each organism: Candida albicans (n = 26), Candida parapsilosis (n = 9) and Candida tropicalis (n = 8).

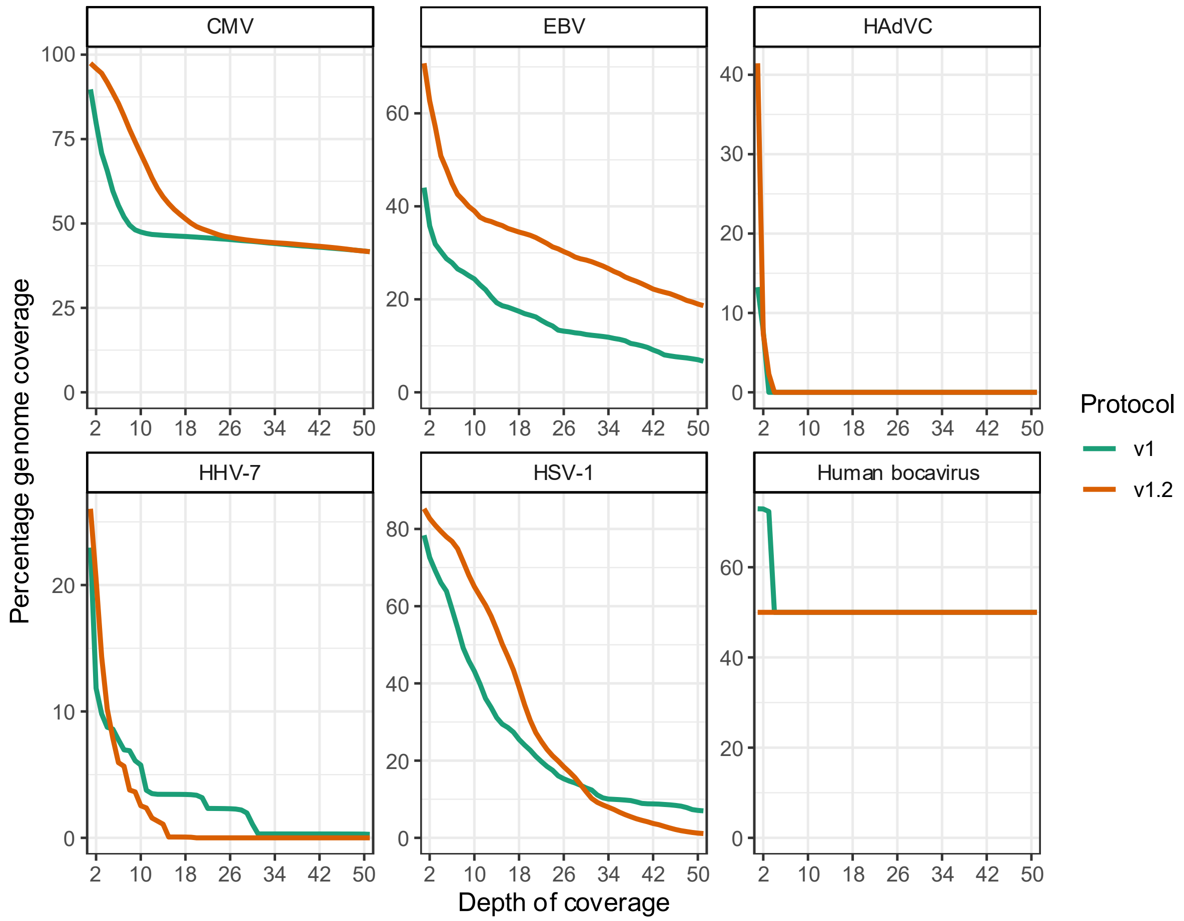

Supplementary figure 14. Median v1 and v1.2 DNA virus genome coverage for individual DNA viruses.

Number of paired samples compared for each organism: CMV (n = 2), EBV (n = 2), HadVC (n = 1), HHV-7 (n = 2), HSV-1 (n=1) and Human bocavirus (n = 2).

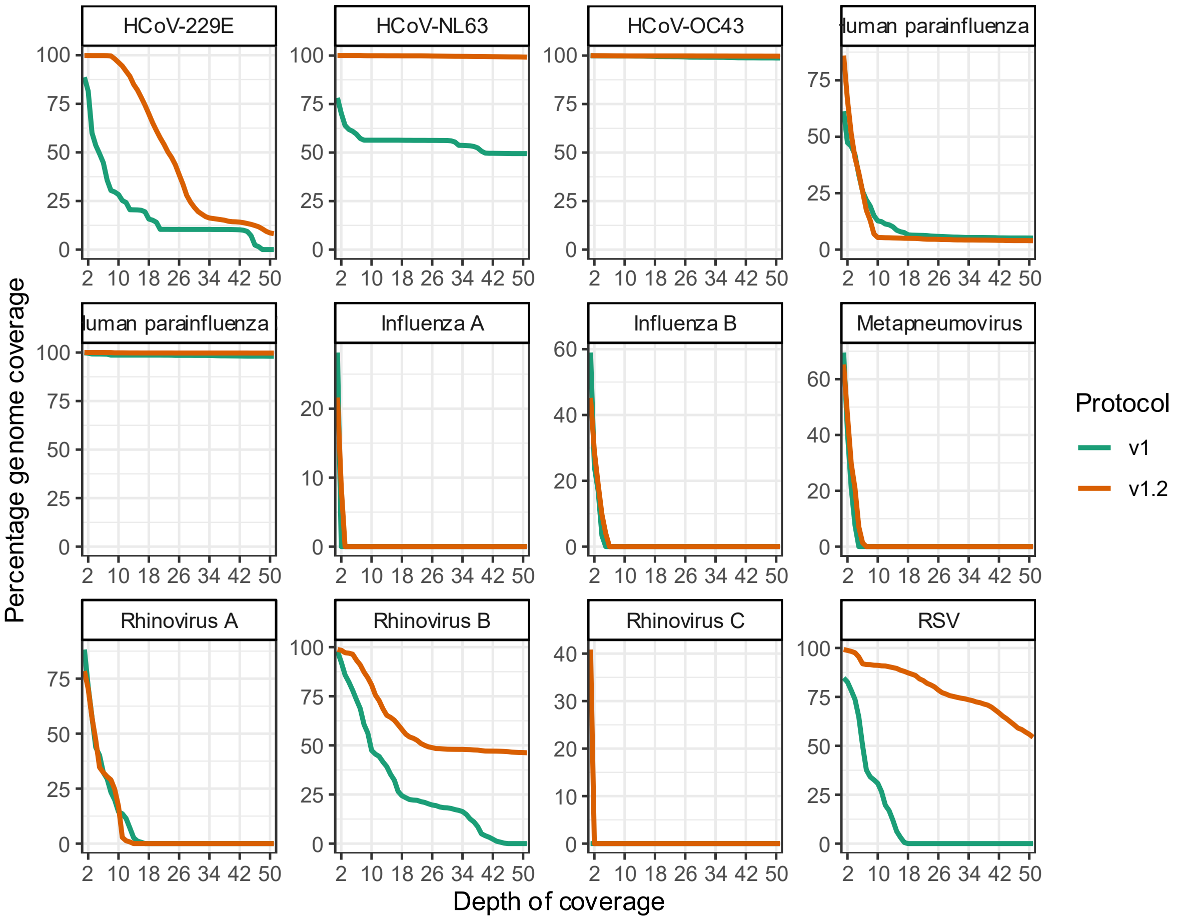

Supplementary figure 15. Median v1 and v1.2 RNA virus genome coverage for individual RNA viruses.

Number of paired samples compared for each organism: HCoV-229E (n = 1), HCoV-NL63 (n = 2), HCoV-OC43 (n = 1), Human parainfluenza 2 (n = 2), Human parainfluenza 3 (n = 1), Influenza A (n = 1), Influenza B (n = 1), Metapneumovirus (n = 3), Rhinovirus A (n = 4), Rhinovirus B (n = 2), Rhinovirus C (n = 1), RSV (n = 5).

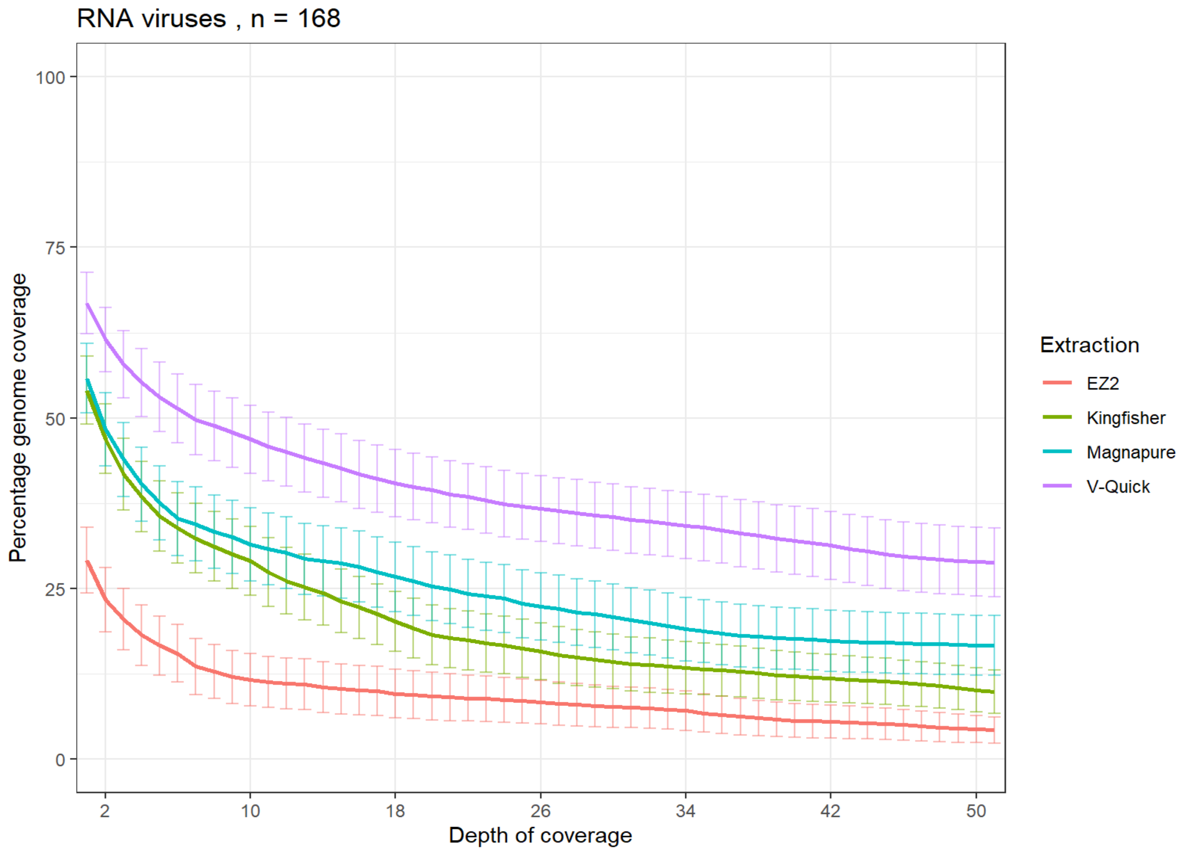

Supplementary figure 16. RNA virus genome coverage of the Zeptometrix Respiratory Panel 2.1 (RP2.1) Control compared by RNA extraction system used.

The lines represent the mean genome coverage and the bars standard error. The analysis included the following organisms: HCoV-229E, HCoV-NL63, HCoV-OC43, Influenza A H1N1, Influenza A H3N2, Influenza B, Metapneumovirus, Human parainfluenza 1, Human parainfluenza 2, Human parainfluenza 3, Human parainfluenza 4, Rhinovirus A, RSV, SARS-CoV-2.

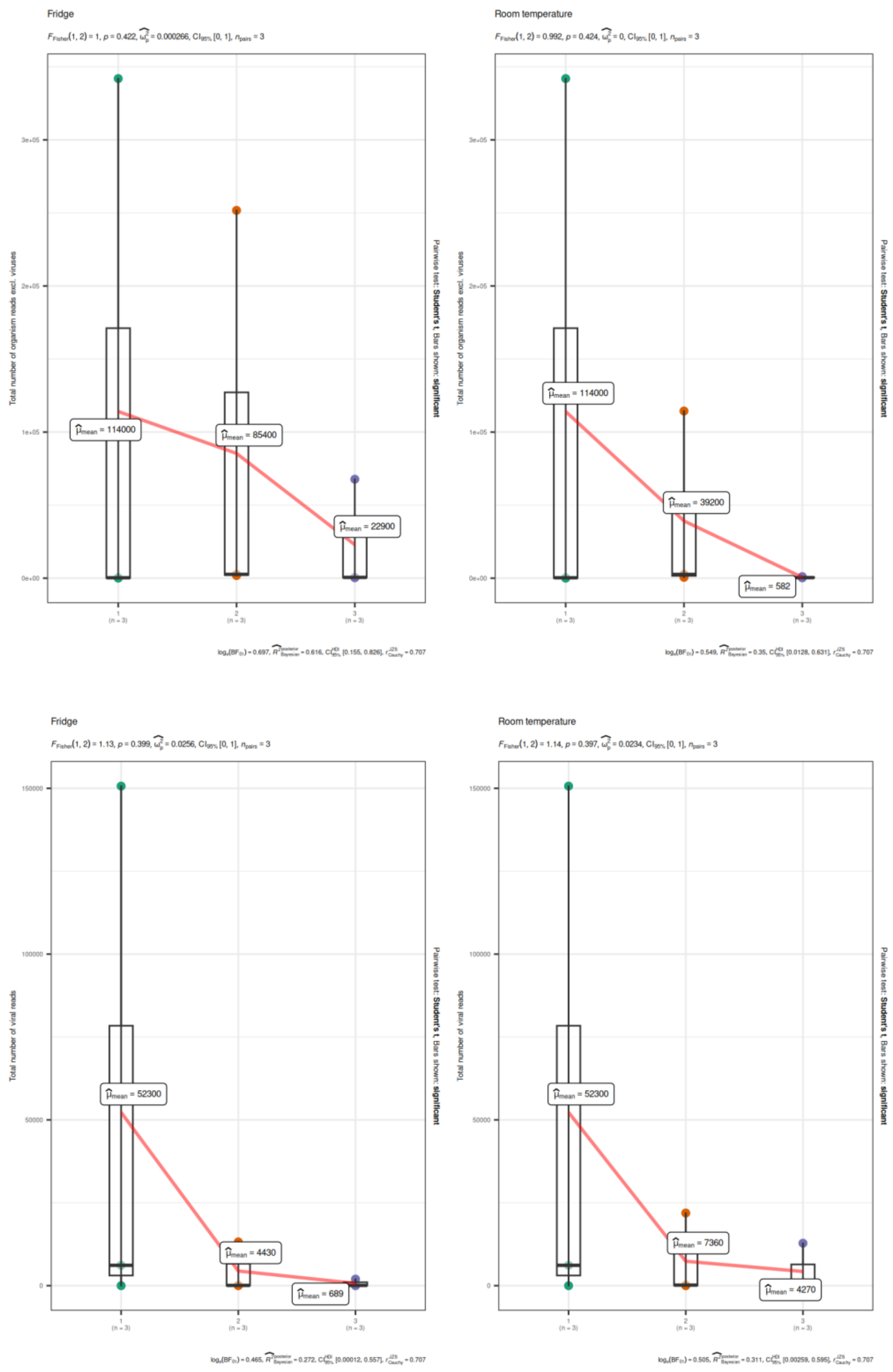

Supplementary figure 17. Comparison of pathogen read recovery from 3 matched clinical samples sequenced on day 1, day 2 and day 3.

This included A) Y-axis presents total reads for all organisms excluding viruses after storage in the fridge vs at room temperature with number of days on x-axis. B) Y-axis presents total reads for all viruses after storage in the fridge and at room temperature with number of days on x-axis.
